# Testing Healthcare Workers Exposed to COVID19 using Rapid Antigen Detection

**DOI:** 10.1101/2020.08.12.20172726

**Authors:** Victor Herrera, Vincent Hsu, Ademola Adewale, Lee Johnson, Timothy Hendrix, Jeffrey Kuhlman, Neil Finkler

## Abstract

There is a need to develop safe and cost-effective ways to test healthcare workers for COVID19. Here we describe a rapid antigen testing strategy in a cohort of 497 Healthcare workers exposed to SARS-CoV-2 that can be applied by systems facing a surge of COVID19 cases, increased number of exposures in their workforce and limited RT-PCR availability. Our findings support an expanded use for antigen testing beyond its current indication and highlights the importance of further evaluating this modality for the diagnosis of COVID19 on asymptomatic individuals.

## Introduction

As cases of COVID19 cases started to rise again in Florida at the beginning of June ^1^, our healthcare system faced multiple challenges due to an increasing number of exposures in our workforce, limited availability of RT-PCR capacity and rapidly increasing patient volume. Consequently, we had to find a rapid solution that was safe and effective to test Health Care Workers (HCWs) exposed to SARS-CoV-2.

## Methods

Our system was an early adopter of rapid antigen testing after this platform received EUA approval by the FDA on May, 2020^2^. Our Ambulatory Centers completed a validation of this modality on 1172 patients that presented with symptoms, by testing all of them with both Antigen Testing and RT-PCR as the gold standard. (Sensitivity 77% 95%CI 72.55% to 80.48% and Specificity 99.1% 95%CI 98.18% to 99.69%) (See Table 1). We used this experience to design an antigen-based testing pilot for HCWs exposed to COVID19 between June 18 and July 21, 2020. A screening algorithm was developed that required exposed HCWs to be tested not sooner than 4-5 days post exposure if no symptoms, or sooner if they became symptomatic. We tested all asymptomatic and symptomatic HCWs in contact with confirmed COVID19 cases at work or in the community. An asymptomatic HCWs that tested Antigen negative was considered a true negative if it was a low-risk exposure, while symptomatic HCWs that tested negative received a confirmatory SARS-CoV-2 PCR test. All High Risk exposures were required to self-isolate at home regardless of results on antigen testing.

**Table 1.** Results of Internal Validation on 1172 patients presenting with COVID19 symptoms to AdventHealth Centracare centers and tested with both SARS-CoV-2 Antigen Testing and SARS-CoV-2 RT-PCR as gold standard. Sensitivity 77% 95%CI 72.55% to 80.48% and Specificity 99.1% 95%CI 98.18% to 99.69%. Patients tested with SARS-Co-V-2 antigen were not limited to those with symptoms onset within 5 days which may have resulted on a lower sensitivity for antigen testing on this validation based on updated recommendations from the manufacturer for optimal time window for testing ^2^.

| *Antigen Pilot (CFD-S)* |  | Coronavirus 2019 PCR Test |  | **76.69%**<br>Sensitivity |
| --- | --- | --- | --- | --- |
|  |  | Detected | Not Detected |  |
| Coronavirus 2019<br>Antigen Test | Detected | **352**<br>(A) True Positive | **6**<br>(B) False Positive | Sensitivity = A / (A+C) |
|  | Not Detected | **107**<br>(C) False Negative | **707**<br>(D) True Negative | **99.16%**<br>Specificity<br>Specificity= D / (B+D) |

AdventHealth

## Results

497 HCWs exposed to COVID19 participated in this pilot (figure 1). Of those tested, 358 were asymptomatic and 139 symptomatic. Of the asymptomatic, 7 were positive (2%) on rapid antigen testing, while 351 (98%) were negative. Of the 139 symptomatic HCWs, 15 (11%) tested positive with antigen, while 124 (89.2%) tested negative. Out of this group, 112 had a confirmatory SARS-CoV-2 RT-PCR with 5 (4.5%) testing positive, and 107 (95.5%) confirmed negative. Overall, 27 (5%) out of 497 exposed HCWs had a positive test for COVID19. HCWs with a positive antigen on RT-PCR were excluded from work, as well as those with no symptoms but a high risk exposure as per CDC guidelines.^2^ Those that had a low risk exposure with no symptoms and had a negative antigen result returned to work. None of the asymptomatic antigen negative HCWs developed symptoms or had evidence of transmitting SARS-CoV-2. Due to the low positivity rate on RT-PCR for this cohort, these samples were run using a pooled methodology which further helped conserve PCR capacity.

**Figure 1.**
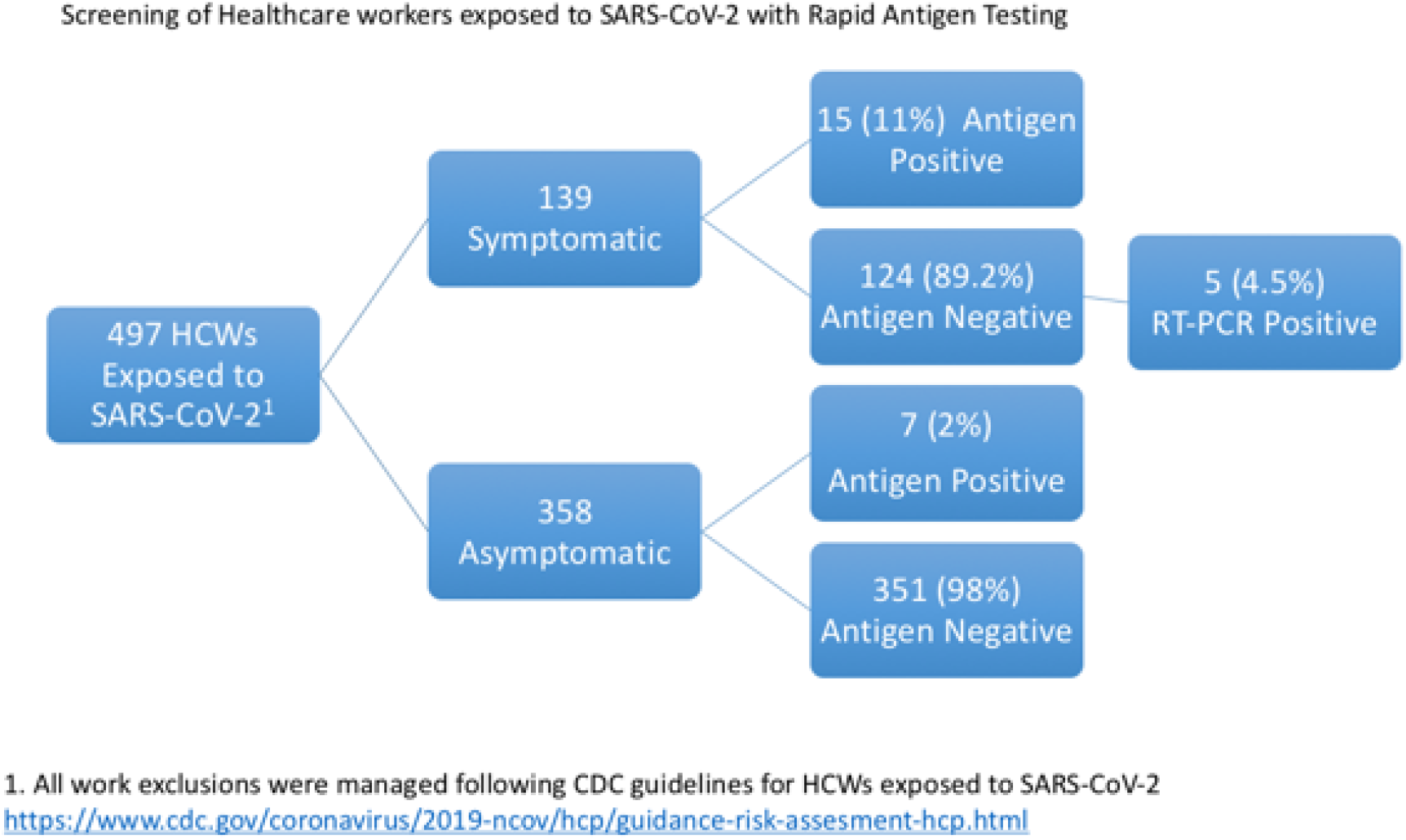

## Conclusions

This testing approach enabled us to rapidly test exposed HCWs and decrease the number of workdays lost with no evidence of an increased risk of COVID19 transmission. Furthermore, this screening allowed us to identify asymptomatic carriers among our HCWs^4^. We believe this finding underscores the importance of considering testing exposed HCWs even if asymptomatic. A potential limitation of this study included not doing a SARS-CoV-2 RT-PCR for HCWs with negative antigen results and no symptoms, although we found no evidence of increased risk of transmission or development of symptoms on this group.

For healthcare systems facing a COVID19 surge and high number of exposures in their workforce, a rapid antigen testing process is an effective strategy that relieves anxiety, decrease resource utilization, and facilitates return to work. It can also serve as a cost effective method to identify asymptomatic HCWs that are SARS-CoV-2 carriers. More data is needed to understand the value of screening asymptomatic cohorts for COVID19 including HCWs, and the potential role of rapid antigen testing for this strategy.

## Data Availability

All data generated or analysed during this study are included in this published article (and its supplementary information files if applicable).

